# Performance management systems among physiotherapists in public rehabilitation centers in Bangladesh: A cross-sectional study of the health workforce

**DOI:** 10.64898/2026.07.09.26357613

**Authors:** Sucheta Kanan, Polok Halder, Abdullah Shuchorit, Md. Hamidur Rahman, Tamal Ghosh Trikta, Tania Islam Liza, Bonosree Rani Borsha, Imrul Kays, Raju Ahmed

**Affiliations:** BRB Hospitals Limited, 77/A, Panthapath, Dhaka-1215, Bangladesh; Dhaka College of Physiotherapy, House-1039, Road-14/A, Adabor Main Road, Dhaka-1207, Bangladesh; Department of Public Health & Informatics, Bangladesh Medical University (BMU), Shahbag, Dhaka-1000, Bangladesh; Protibondhi Seba O Sahajjo Kandro (PSOSK) - Dhamrai, Jatiyo Protibondhi Unnayan Foundation, Ministry of Social Welfare, Dhaka-1350, Bangladesh

**Keywords:** performance management, physiotherapist, rehabilitation, health workforce, Bangladesh, public health

## Abstract

Health workforce performance is central to service quality, yet little empirical work has examined how performance management systems operate for physiotherapists in rehabilitation services in low- and middle-income settings. This cross-sectional study assessed the current state, perceived effectiveness, and process gaps of performance management systems among physiotherapists working in public rehabilitation centers in Dhaka, Bangladesh. A pretested semi-structured questionnaire was administered to 105 physiotherapists between September and October 2025. Descriptive statistics were used to summarize participant characteristics and performance management indicators. Wilson 95% confidence intervals were estimated for key proportions. A nine-item exploratory performance management system maturity score was constructed from process indicators. Fisher exact tests with Cramer’s V were used to examine associations with perceived system effectiveness, and exploratory logistic regression estimated odds ratios for effective or moderately effective performance management. The mean age of respondents was 31.6 years, 56 of 105 were male, and 85 of 105 had graduate or postgraduate qualifications. Formal performance management systems were reported by 102 of 105 respondents (97.1%, 95% CI 91.9-99.0). Standardized appraisal timing and method, assessment form use, performance planning, and formal evaluation systems were each reported by about 60-70% of participants. Reward-performance linkage was perceived as motivating by 97 of 105 respondents (92.4%, 95% CI 85.7-96.1). Overall, 81 of 105 respondents (77.1%, 95% CI 68.2-84.1) rated the system as effective or moderately effective. Training recipient category was associated with perceived effectiveness (Fisher exact p=0.0035; Cramer’s V=0.363), as was perceived appropriateness of the process (p=0.0323; Cramer’s V=0.258). The maturity score was not independently associated with perceived effectiveness in exploratory regression. Public rehabilitation centers in Dhaka appear to have formal performance management systems, but the systems are only moderately developed. Strengthening training coverage, transparent evaluation criteria, routine feedback, and formal system review may improve staff confidence in performance management processes.

## Introduction

A well-performing health workforce is a core requirement for universal health coverage and quality health services. Global health policy has placed increasing emphasis on health workforce availability, performance, accountability, and supportive management systems [1,2]. At the same time, the demand for rehabilitation is rising rapidly. Global estimates suggest that more than one-third of the world population may need rehabilitation at some point in life [3]. The World Health Organization has therefore urged countries to strengthen rehabilitation leadership, planning, governance, and workforce capacity as part of health system strengthening [4].

These issues are highly relevant in Bangladesh. Rehabilitation services for persons with disabilities remain unevenly distributed and insufficiently integrated into mainstream health services [5]. The physiotherapy profession in Bangladesh has also faced historical challenges related to recognition, access, and workforce development [6]. Improving the management of physiotherapists is therefore not only an internal organizational issue but also a health-system concern linked with service readiness, quality of care, and rehabilitation access.

Performance management is broader than annual performance appraisal. It involves defining expectations, aligning individual and organizational goals, monitoring performance, providing feedback, supporting development, and using information for fair decisions [7–10]. In public-sector organizations, performance systems can support accountability and learning when used appropriately, but they may also become administrative exercises if evaluation criteria are unclear or feedback is weak [11]. Employee perceptions are particularly important because the acceptance, fairness, and usefulness of the system influence how staff engage with the process [12].

In physiotherapy, much of the existing literature focuses on clinical competence, professional identity, evidence-based practice, and job performance rather than the organizational systems used to manage performance [13,14]. Evidence from Bangladesh on performance management among physiotherapists is especially limited. This study aimed to assess the current state, process characteristics, perceived effectiveness, and key gaps of performance management systems among physiotherapists in public rehabilitation centers in Dhaka, Bangladesh.

## Materials and methods

### Study design and reporting

This was a cross-sectional health workforce study reported in line with the STROBE guidance for observational studies [15]. The study assessed physiotherapists’ self-reported experience and perceptions of performance management systems in public rehabilitation settings.

### Study setting and period

The study was conducted in public rehabilitation centers located in Dhaka, Bangladesh, including Dhaka Medical College Hospital, Shaheed Suhrawardy Medical College Hospital, the National Institute of Traumatic and Orthopedic Rehabilitation, and the Centre for the Rehabilitation of the Paralysed. Data collection was conducted from 10 September to 15 October 2025 within a broader study period from January to December 2025.

### Participants and eligibility criteria

The study population consisted of physiotherapists working in the selected rehabilitation centers. Eligible participants were male or female physiotherapists who had worked for more than one year and provided written informed consent. Physiotherapists who did not provide written informed consent were not enrolled. A convenience sampling approach was used because participants were recruited according to availability within the selected centers. Recruitment was conducted through convenience sampling within the selected rehabilitation centres. The numbers of physiotherapists approached, assessed for eligibility, and declining participation were not recorded prospectively. Consequently, reasons for non-participation were unavailable. The final analytic dataset comprised 105 respondents.

### Sample size

The sample size was calculated using n=z2pq/d2 with z=1.96, p=0.36, q=0.64, and d=0.05. The initial sample size was 354. Applying finite population correction for an estimated population of 150 physiotherapists yielded a required sample of approximately 105 participants. The final analytic dataset included 105 respondents.

### Data collection tool and variables

Data were collected using a pretested semi-structured questionnaire developed from study objectives and relevant performance management literature. The questionnaire covered socio-demographic characteristics, work profile, current performance management system, appraisal methods, perceived challenges, performance planning, documentation, feedback, training, bias reduction, reward linkage, evaluation practices, perceived effectiveness, and planned system changes. A 14-item ability/performance rating section captured domains such as work quality, knowledge, adaptation ability, judgment, attendance/punctuality, leadership ability, planning, and communication ability.

### Data cleaning and derived variables

The dataset was cleaned before analysis. Duplicate respondent IDs were checked, invalid or inconsistent entries were reviewed, and skip-pattern items were set to missing when the parent item indicated that the question was not applicable. Service duration was converted to years using conservative rules based on raw entries. Perceived effectiveness was dichotomized as effective or moderately effective versus ineffective or don’t know. An exploratory performance management system maturity score ranging from 0 to 9 was calculated by summing nine binary indicators: formal system present, assessment form used, process perceived as appropriate, overall rating given, performance planning undertaken, three or more meetings per year, process carried out objectively, reward-performance link perceived as motivating, and formal evaluation system present. The score was not treated as a validated scale; it was used as a process completeness indicator.

### Statistical analysis

Categorical variables were summarized as frequencies and percentages. Continuous variables were summarized using mean and standard deviation, and median with interquartile range. Wilson 95% confidence intervals were calculated for key proportions [16]. Internal consistency of the 14-item rating section was assessed using Cronbach’s alpha [17]. Fisher exact tests were used to examine associations between selected categorical predictors and perceived effectiveness; simulated p-values were used where table dimensions required. Cramer’s V was reported as an effect size for categorical associations. Exploratory logistic regression estimated odds ratios and 95% confidence intervals for perceived effectiveness. Statistical significance was set at alpha=0.05. Because the inferential analyses were exploratory and the study was not powered for multiple hypothesis testing, p-values were interpreted cautiously and no multiple-comparison correction was applied. Analyses were performed in R using the tidyverse package [18,19] and standard categorical data methods [20].

### Ethics statement

The study protocol was approved by the Institutional Review Board of the National Institute of Preventive and Social Medicine, Bangladesh (Memo no. NIPSOM/IRB/2023/06; approval date: 02 July 2023). All participants provided written informed consent before data collection. The analytic dataset was anonymized before analysis.

## Results

### Participant characteristics

The final dataset included 105 physiotherapists. The mean age was 31.6 years (SD 5.6), and 61 participants (58.1%) were aged 21-30 years. Male physiotherapists accounted for 56 of 105 respondents (53.3%). Most respondents had graduate or postgraduate qualifications (85 of 105, 81.0%), and 60 of 105 (57.1%) worked in physical medicine departments. The median service duration was 4.0 years (interquartile range 2.5-6.0). Table 1 summarizes participant characteristics and work profile.

**Table 1.** Participant characteristics and work profile (n=105).

| Variable | Category | n (%) or summary |
| --- | --- | --- |
| Age, years |  | 31.62 (5.57) |
| Monthly income, BDT |  | 46509.52 (26084.18) |
| Service duration, years |  | 5.72 (5.18) |
| Mean 14-item rating score |  | 2.88 (0.88) |
| PMS maturity score, 0-9 |  | 6.08 (1.73) |
| Sex | Female | 49 (46.7%) |
| Sex | Male | 56 (53.3%) |
| Age group | 21-30 years | 61 (58.1%) |
| Age group | 31-40 years | 36 (34.3%) |
| Age group | 41-50 years | 5 (4.8%) |
| Age group | 51-60 years | 3 (2.9%) |
| Educational qualification | Diploma | 20 (19%) |
| Educational qualification | Graduate | 44 (41.9%) |
| Educational qualification | Postgraduate | 41 (39%) |
| Monthly income | 30,000-39,000 BDT | 43 (41%) |
| Monthly income | 40,000-50,000 BDT | 22 (21%) |
| Monthly income | <30,000 BDT | 10 (9.5%) |
| Monthly income | >50,000 BDT | 30 (28.6%) |
| Marital status | Married | 88 (83.8%) |
| Marital status | Unmarried | 17 (16.2%) |
| Type of family | Extended family | 35 (33.3%) |
| Type of family | Nuclear family | 70 (66.7%) |
| Area of residence | Outside hospital campus | 104 (99%) |
| Area of residence | Within hospital campus | 1 (1%) |
| Working department | Musculoskeletal | 20 (19%) |
| Working department | Neurology | 12 (11.4%) |
| Working department | Pediatric | 11 (10.5%) |
| Working department | Physical Medicine | 60 (57.1%) |
| Working department | Prosthesis and Orthosis | 2 (1.9%) |
| Service duration | 1-5 years | 73 (70.9%) |
| Service duration | 11-15 years | 9 (8.7%) |
| Service duration | 16+ years | 3 (2.9%) |
| Service duration | 6-10 years | 17 (16.5%) |
| Service duration | <1 year | 1 (1%) |
| Working days per week | 5 days/week | 21 (20%) |
| Working days per week | 6 days/week | 84 (80%) |
| Shift | Both morning and evening | 20 (19%) |
| Shift | Evening | 6 (5.7%) |
| Shift | Morning | 79 (75.2%) |

*Continuous variables are reported as mean (SD). Categorical variables are reported as n (%). Service* *duration had two missing category values after cleaning.*

### Performance management system indicators

Formal performance management systems were reported by 102 of 105 respondents (97.1%, 95% CI 91.9-99.0). Standardized appraisal at the same time and by the same method was reported by 73 of 105 respondents (69.5%, 95% CI 60.2-77.5), and assessment form use was also reported by 73 of 105 respondents. About three-fifths reported that overall ratings were given, performance planning was undertaken, and at least three meetings per year were scheduled. Reward linkage was perceived as motivating by 97 of 105 respondents (92.4%, 95% CI 85.7-96.1). Overall, 81 of 105 respondents (77.1%, 95% CI 68.2-84.1) rated the performance management process as effective or moderately effective. Table 2 and Fig 1 summarize key performance management indicators.

**Fig 1.**
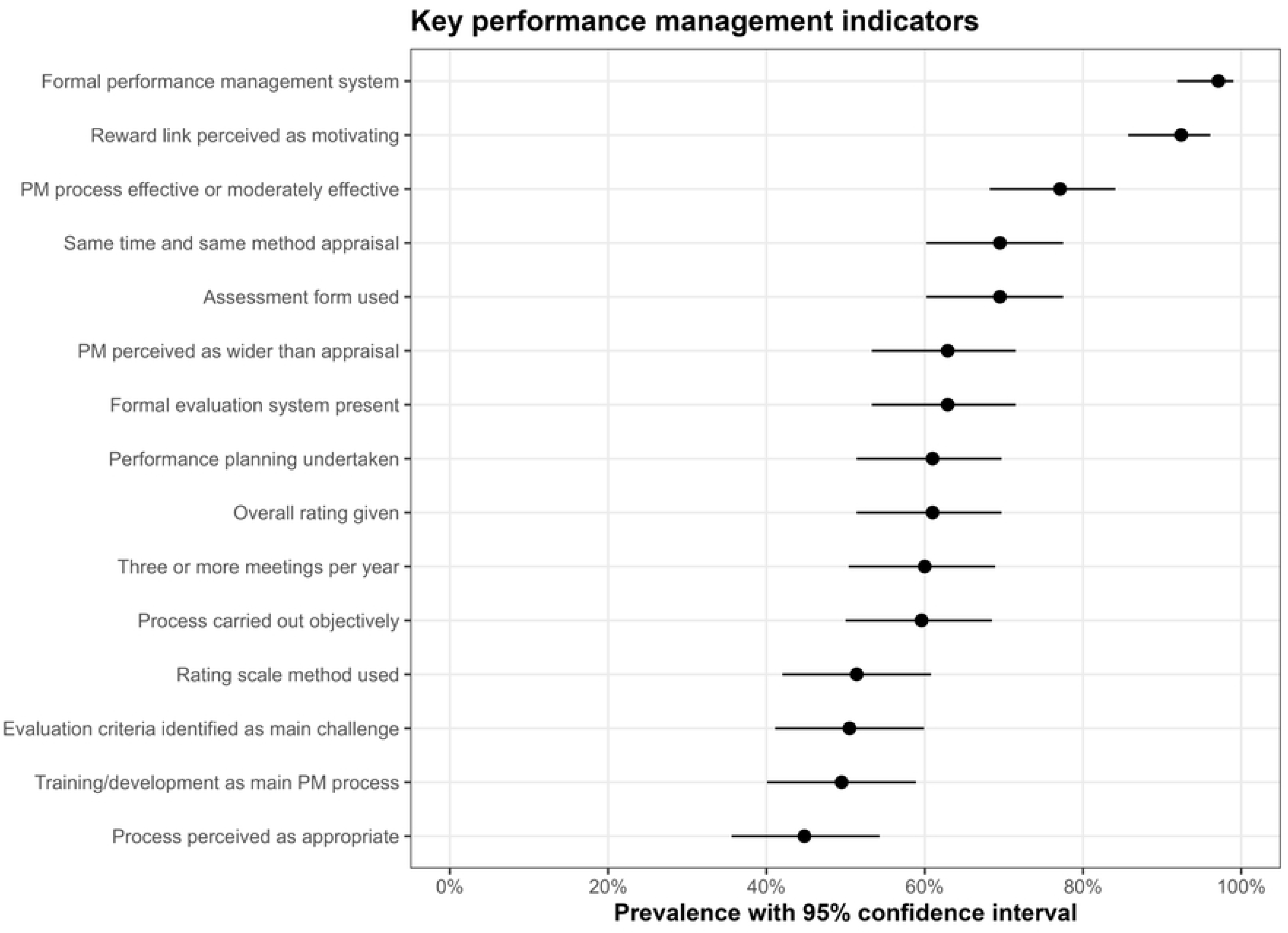
Key performance management indicators with 95% confidence intervals. The figure shows the prevalence of selected performance management system indicators among 105 physiotherapists. Confidence intervals were calculated using the Wilson method.

**Table 2.** Key performance management indicators with Wilson 95% confidence intervals.

| Indicator | n/N | Percent (95% CI) |
| --- | --- | --- |
| Formal performance management system | 102/105 | 97.1 (91.9-99) |
| Same time and same method appraisal | 73/105 | 69.5 (60.2-77.5) |
| Assessment form used | 73/105 | 69.5 (60.2-77.5) |
| Rating scale method used | 54/105 | 51.4 (42-60.8) |
| Training/development as main PM process | 52/105 | 49.5 (40.1-58.9) |
| Evaluation criteria identified as main challenge | 53/105 | 50.5 (41.1-59.9) |
| PM perceived as wider than appraisal | 66/105 | 62.9 (53.3-71.5) |
| Process perceived as appropriate | 47/105 | 44.8 (35.6-54.3) |
| Overall rating given | 64/105 | 61 (51.4-69.7) |
| Performance planning undertaken | 64/105 | 61 (51.4-69.7) |
| Three or more meetings per year | 63/105 | 60 (50.4-68.9) |
| Process carried out objectively | 62/104 | 59.6 (50-68.5) |
| Reward link perceived as motivating | 97/105 | 92.4 (85.7-96.1) |
| Formal evaluation system present | 66/105 | 62.9 (53.3-71.5) |
| PM process effective or moderately effective | 81/105 | 77.1 (68.2-84.1) |

*PM, performance management.*

### Performance management maturity and rating profile

The exploratory performance management system maturity score had a mean of 6.08 (SD 1.73) on a 0-9 scale, with a median of 6.0 (interquartile range 5.0-7.0). The score distribution suggested moderate system completeness across the sample (Fig 2). The 14-item ability/performance rating section showed high internal consistency (Cronbach’s alpha=0.954; complete cases=105). Mean item scores ranged from 2.60 for self-work knowledge to 3.23 for attendance/punctuality (Fig 3).

**Fig 2.**
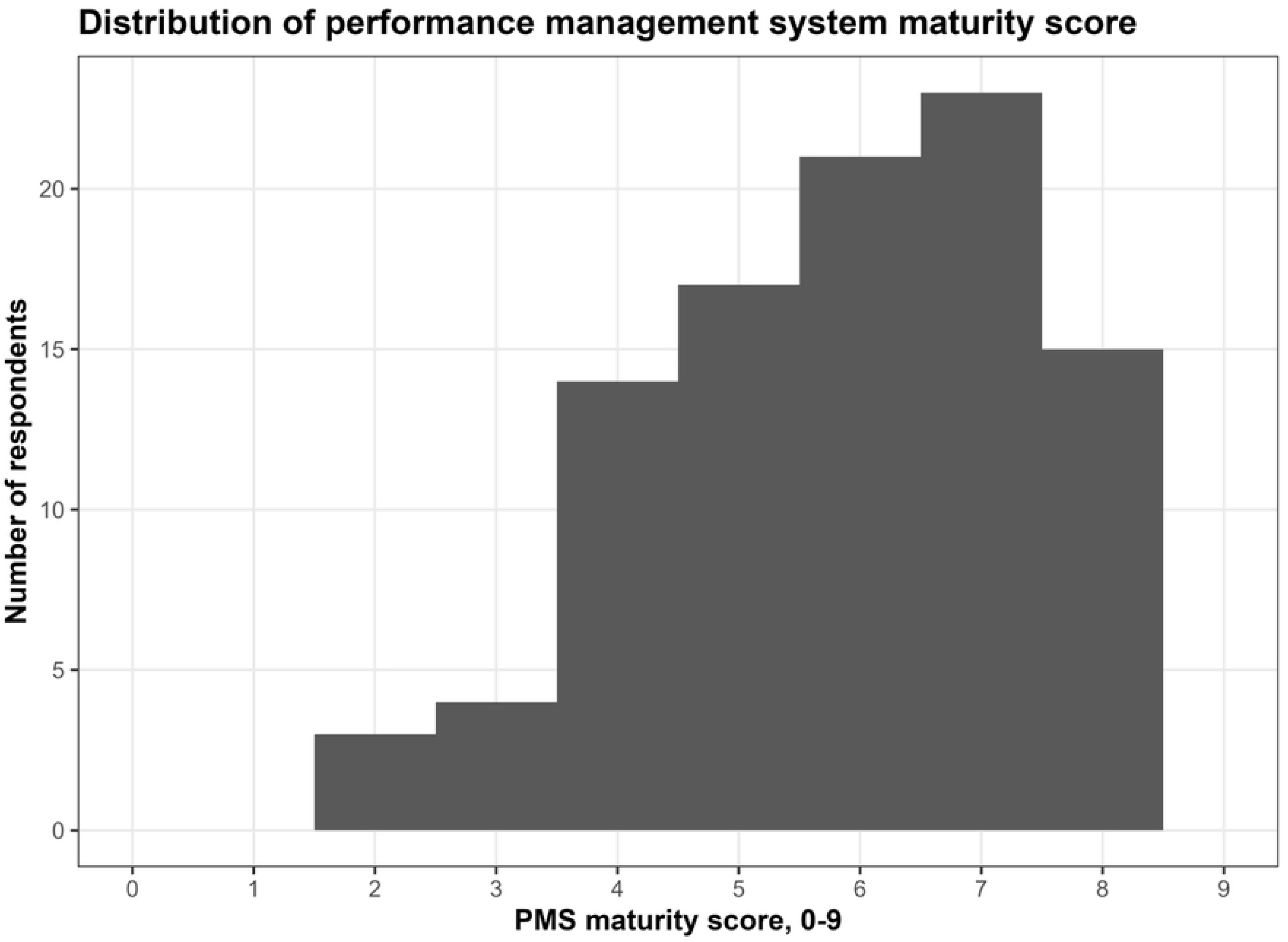
Distribution of the performance management system maturity score. The exploratory score ranged from 0 to 9 and summarized nine process indicators. Higher values indicate more complete reported performance management processes.

**Fig 3.**
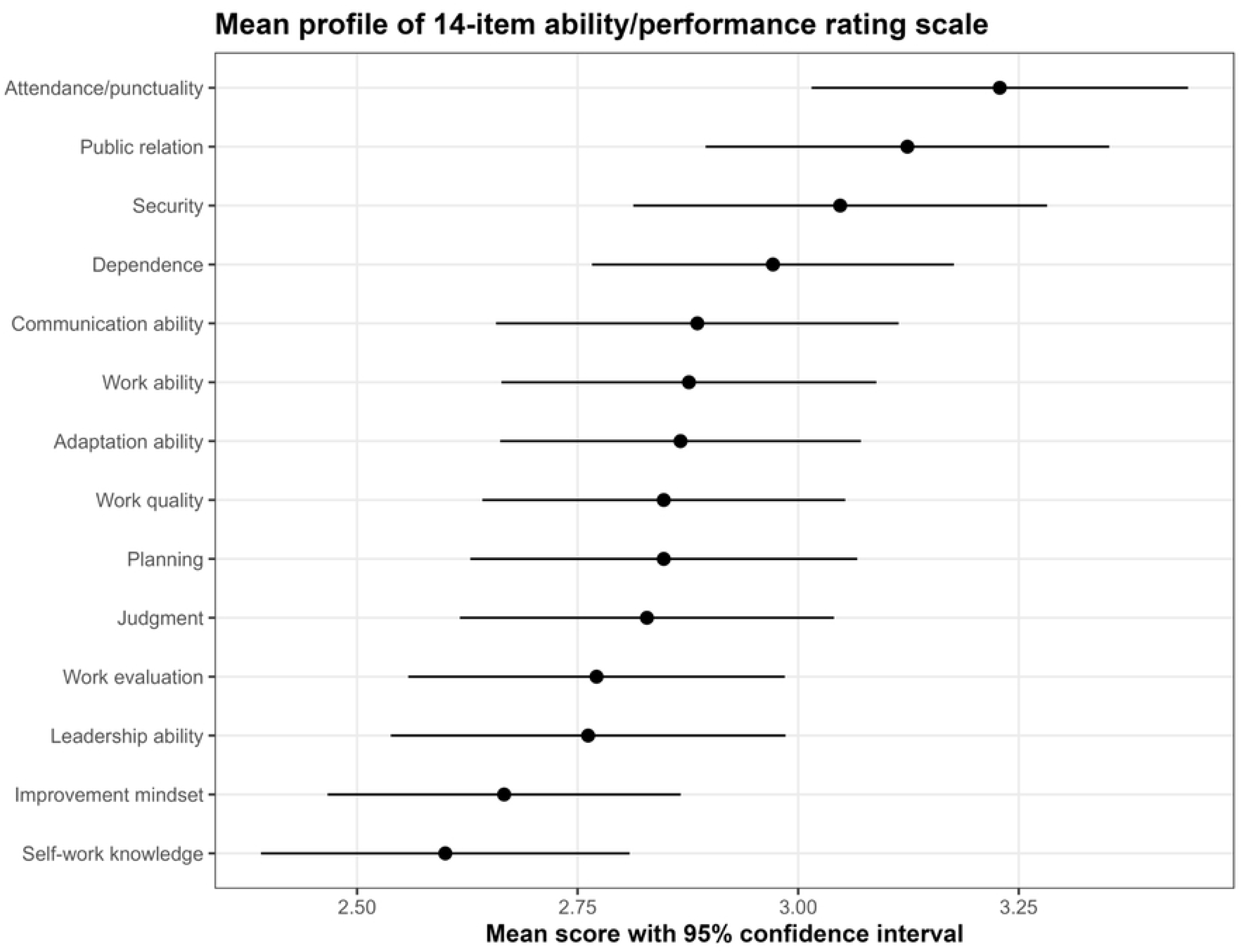
Mean profile of the 14-item ability/performance rating section. Points indicate item means and horizontal bars indicate 95% confidence intervals. The item set showed high internal consistency, but the score should be interpreted as exploratory because the tool was locally developed.

### Associations with perceived effectiveness

Two factors were associated with perceived effectiveness. Training recipient category showed a moderate association with perceived effectiveness (Fisher exact p=0.0035; Cramer’s V=0.363). Perceived appropriateness of the performance management process also showed a small-to-moderate association with effectiveness (p=0.0323; Cramer’s V=0.258). Most demographic variables, formal system presence, rating use, performance planning, meeting frequency, formal evaluation, and maturity category were not statistically associated with perceived effectiveness (Table 3 and Fig 4).

**Fig 4.**
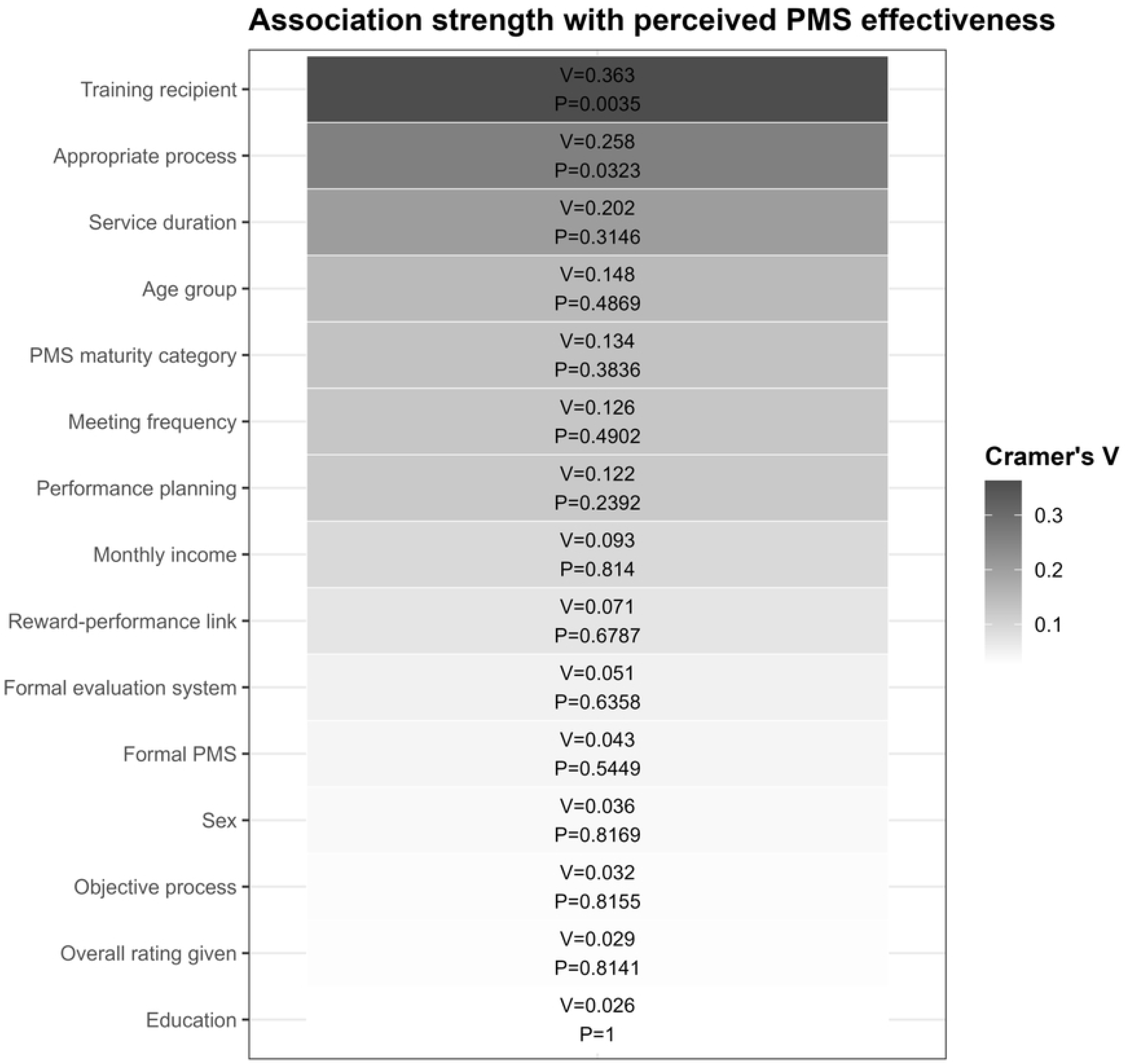
Association strength with perceived performance management system effectiveness. The heatmap summarizes Fisher exact test p-values and Cramer’s V effect sizes for selected predictors.

**Table 3.** Associations between selected variables and perceived performance management effectiveness.

| Variable | N | P value | Cramer's V |
| --- | --- | --- | --- |
| Sex | 105 | 0.8169 | 0.036 |
| Age group | 105 | 0.4869 | 0.148 |
| Education | 105 | 1.0 | 0.026 |
| Monthly income | 105 | 0.814 | 0.093 |
| Working department | 105 | 0.2133 | Not estimable |
| Service duration | 103 | 0.3146 | 0.202 |
| Formal PMS | 105 | 0.5449 | 0.043 |
| Appropriate process | 105 | 0.0323 | 0.258 |
| Overall rating given | 105 | 0.8141 | 0.029 |
| Performance planning | 105 | 0.2392 | 0.122 |
| Meeting frequency | 105 | 0.4902 | 0.126 |
| Training recipient | 105 | 0.0035 | 0.363 |
| Objective process | 104 | 0.8155 | 0.032 |
| Reward-performance link | 105 | 0.6787 | 0.071 |
| Formal evaluation system | 105 | 0.6358 | 0.051 |
| PMS maturity category | 105 | 0.3836 | 0.134 |

*Outcome was effective/moderately effective versus ineffective/don't know. P-values are from Fisher exact tests. Cramer's V values around 0.1, 0.3, and 0.5 may be interpreted as small, moderate, and large association strength in broad terms, but interpretation depends on table dimensions.*

In exploratory logistic regression, the maturity score was not associated with perceived effectiveness in the unadjusted model (OR 1.11, 95% CI 0.85-1.45; p=0.4329) or after adjustment for mean rating score and age (OR 1.15, 95% CI 0.88-1.51; p=0.3052). The mean 14-item rating score and age were also not independently associated with perceived effectiveness (Table 4).

**Table 4.** Exploratory logistic regression for perceived performance management effectiveness.

| Model | Term | OR (95% CI) | P value |
| --- | --- | --- | --- |
| Model 1: unadjusted | PMS maturity score, per 1-point increase | 1.11 (0.85-1.45) | 0.4329 |
| Model 2: adjusted | PMS maturity score, per 1-point increase | 1.15 (0.88-1.51) | 0.3052 |
| Model 2: adjusted | Mean 14-item rating score, per 1-point increase | 0.92 (0.51-1.65) | 0.7818 |
| Model 2: adjusted | Age, per 1-year increase | 1.09 (0.96-1.24) | 0.1748 |

*Outcome was effective/moderately effective versus ineffective/don't know. OR, odds ratio; CI, confidence interval; PMS, performance management system.*

## Discussion

### Principal findings

This study found that formal performance management systems were widely reported among physiotherapists in public rehabilitation centers in Dhaka, Bangladesh. However, the presence of a formal system did not mean that all core processes were strong. Several process indicators, including overall rating, performance planning, meeting frequency, formal evaluation, and perceived appropriateness, were present in only about 45-63% of respondents. This pattern suggests that performance management exists structurally but remains uneven in implementation.

The most actionable findings relate to training and perceived appropriateness. Training recipient category had the strongest association with perceived effectiveness, and respondents who perceived the system as appropriate were more likely to rate it as effective or moderately effective. These findings are consistent with performance management theory, which emphasizes that performance systems require supervisor capability, employee understanding, clear evaluation criteria, and developmental feedback rather than documentation alone [7–12].

The high proportion of respondents who perceived reward linkage as motivating suggests that physiotherapists value recognition and fair linkage between performance and career or financial outcomes. This does not mean that reward linkage alone should drive the system. Public-sector performance measurement research cautions that poorly designed incentive systems may distort behavior if goals and indicators are unclear [11]. In this study, half of respondents identified evaluation criteria as a main challenge, suggesting that transparency and fairness may be more urgent than adding reward mechanisms without system strengthening.

The 14-item rating section had high internal consistency. This may indicate that the items measured a common perception of performance-related ability, but a Cronbach’s alpha above 0.95 can also indicate redundancy among items. The rating section should therefore be interpreted cautiously. Further psychometric work would be needed before using this tool as a formal validated scale.

The exploratory maturity score was not associated with perceived effectiveness in regression. This finding has two possible explanations. First, process presence may not be enough; how the process is implemented may matter more than whether a component exists. Second, the sample size was modest and the score was newly constructed, so the analysis may have had limited ability to detect small associations.

The regression results should not be interpreted as evidence that maturity is irrelevant; rather, they show that this specific exploratory score did not independently explain perceived effectiveness in this dataset.

### Relevance to rehabilitation and health systems

The findings have practical relevance for rehabilitation workforce management in Bangladesh. Rehabilitation services depend on skilled professionals whose performance is influenced by supervision, feedback, training, and organizational support. Bangladesh has documented rehabilitation service gaps and workforce constraints [5,6].

Strengthening performance management may support service quality by improving role clarity, routine feedback, and staff development, especially in public facilities where rehabilitation demand is likely to increase.

### Implications for policy and management

Public rehabilitation centers should move from appraisal-oriented documentation toward developmental performance management. Practical steps include defining transparent evaluation criteria for physiotherapy roles, training line managers and senior physiotherapists in feedback and appraisal skills, scheduling mid-year and annual review meetings, documenting performance plans, creating a fair process for identifying training needs, and using performance data for service improvement rather than only for promotion or administrative decisions. Facility managers should also evaluate whether staff understand the system and whether evaluation criteria are perceived as fair.

### Strengths and limitations

This study provides one of the first structured assessments of performance management systems among physiotherapists in public rehabilitation settings in Bangladesh. It used a cleaned anonymized dataset, confidence intervals for key proportions, reliability assessment of the rating section, effect size estimates for categorical associations, and exploratory regression. The study also has limitations. The cross-sectional design prevents causal inference. Convenience sampling may have introduced selection bias, and the sample was limited to selected centers in Dhaka. Because recruitment-stage counts and reasons for non-participation were not recorded, the extent and direction of potential non-response bias could not be quantified. Data were self-reported and may be affected by social desirability or recall bias. The maturity score and rating section were exploratory and should not be treated as externally validated instruments. Finally, because some performance management components had a very high prevalence, the study had limited variability for some association analyses.

## Conclusion

Formal performance management systems were widely reported among physiotherapists in public rehabilitation centers in Dhaka, but implementation appeared only moderately developed. Training coverage, perceived appropriateness of the process, clarity of evaluation criteria, routine feedback, and formal review mechanisms emerged as key areas for improvement. For public rehabilitation services in Bangladesh, strengthening performance management should be treated as a health workforce development issue rather than a narrow administrative exercise. Future research should use larger multi-district samples, center-level comparisons, and validated measures to examine how performance management quality relates to staff outcomes and rehabilitation service quality.

## Data Availability

The de-identified minimal dataset underlying the findings of this study is included as Supporting Information (S1 Data). The R code used for data cleaning, statistical analysis, and figure generation is included as Supporting Information (S1 Code).

## Acknowledgments

The authors thank the participating physiotherapists and the authorities of the participating rehabilitation centers for their cooperation during data collection.

## Supporting information captions

S1 Data. Deidentified analysis dataset used for the study.

S1 Table. Full descriptive table of performance management system variables.

S2 Table. Item-total correlations for the 14-item ability/performance rating section.

S3 Table. Perceived effectiveness by performance management system maturity category.

S1 Code. R analysis code used to generate the study tables and figures.

S1 Checklist. STROBE checklist for the cross-sectional study.

